# Prognostic accuracy of triage tools for adults with suspected COVID-19 in a pre-hospital setting: an observational cohort study

**DOI:** 10.1101/2021.07.27.21261031

**Authors:** Carl Marincowitz, Laura Sutton, Tony Stone, Richard Pilbery, Richard Campbell, Benjamin Thomas, Janette Turner, Peter A. Bath, Fiona Bell, Katie Biggs, Madina Hasan, Frank Hopfgartner, Suvodeep Mazumdar, Jennifer Petrie, Steve Goodacre

## Abstract

**Study Objective:** Tools proposed to triage patient acuity in COVID-19 infection have only been validated in hospital populations. We estimated the accuracy of five risk-stratification tools recommended to predict severe illness and compare accuracy to existing clinical decision-making in a pre-hospital setting.

**Methods:** An observational cohort study using linked ambulance service data for patients attended by EMS crews in the Yorkshire and Humber region of England between 18th March 2020 and 29th June 2020 was conducted to assess performance of the PRIEST tool, NEWS2, the WHO algorithm, CRB-65 and PMEWS in patients with suspected COVID-19 infection. The primary outcome was death or need for organ support.

**Results:** Of 7549 patients in our cohort, 17.6% (95% CI:16.8% to 18.5%) experienced the primary outcome. The NEWS2, PMEWS, PRIEST tool and WHO algorithm identified patients at risk of adverse outcomes with a high sensitivity (>0.95) and specificity ranging from 0.3 (NEWS2) to 0.41 (PRIEST tool). The high sensitivity of NEWS2 and PMEWS was achieved by using lower thresholds than previously recommended. On index assessment, 65% of patients were transported to hospital and EMS decision to transfer patients achieved a sensitivity of 0.84 (95% CI 0.83 to 0.85) and specificity of 0.39 (95% CI 0.39 to 0.40).

**Conclusion:** Use of NEWS2, PMEWS, PRIEST tool and WHO algorithm could improve sensitivity of EMS triage of patients with suspected COVID-19 infection. Use of the PRIEST tool would improve sensitivity of triage without increasing the number of patients conveyed to hospital.

## Background

Emergency Medical Service (EMS) and other urgent and emergency care practitioners assessing patients with suspected COVID-19 infection in the community, must rapidly determine whether patients need treatment in hospital or can safely remain at home. The overall risk of mortality in patients with confirmed infection is around 1% and if conveyance is too liberal, hospitals could be overwhelmed by patients who require no specific treatment.^1^ However, failing to identify a patient at risk of serious deterioration could lead to avoidable harm.^2^

Prognostic research has almost exclusively been conducted in hospital settings and current national and international guidelines for risk-stratification of patients with suspected COVID-19 in the community are consensus based.^1, 3-5^ Clinical acuity scores, such as the UK Royal College of Physicians Early Warning Score, version 2 (NEWS2), have been suggested in some guidelines as a way to risk-stratify patients with suspected COVID-19 infection in the community.^6^ The WHO decision-making algorithm for respiratory infection and CRB-65 are used to risk-stratify patients with bacterial pneumonia and PMEWS for use in patients with influenza.^7-9^ However, the accuracy of these risk-stratification tools has only been validated in hospitalised or non-COVID populations.

NEWS2 has shown good prediction of adverse outcome in patients attending the Emergency Department (ED) with suspected COVID-19.^7^ The PRIEST tool was derived by adding age, sex, and performance status to NEWS2, and validation showed improved prediction compared to NEWS2 alone.^10, 11^ Validation of the PRIEST tool, NEWS2 and other clinical risk-stratification tools recommended for use in hospital in a community setting,^7, 10, 12-14^ could identify the most accurate means to triage need for hospitalisation, thereby reducing unnecessary hospital attendances and improve the identification of those most at risk of serious adverse outcomes.

Our study aimed to:

1. Estimate the accuracy of risk-stratification tools recommended to predict severe illness in adults with suspected COVID-19 infection in a pre-hospital setting.
2. Compare the accuracy of risk-stratification tools to existing clinical decision-making around transport to hospital.

## Methods

### Study Design

This observational cohort study used linked routinely collected EMS data to assess the accuracy in a community setting of five clinical risk-stratification tools (PRIEST tool, NEWS2, WHO algorithm, CRB-65 and PMEWS) recommended for use in hospitalised patients with COVID-19 or similar respiratory infections (triage tools shown in Supplementary Material 1).^7, 10, 12-14^

### Setting

Patients with suspected COVID-19 infection attended by emergency medical services provided by Yorkshire Ambulance Service NHS Trust (YAS). Emergency services provided by YAS covers a region in the north of England of approximately 6,000 square miles and with a population of 5.3 million.

### Data Sources and linkage

EMS providers complete an electronic patient report form (ePRF) each time they attend an emergency call, which records presenting patient characteristics and clinical care in a standardised manner. YAS provided a dataset of ePRF data for all EMS responses between the 18th March 2020 and 29th June 2020 where the attending ambulance staff recorded a clinical impression of suspected or confirmed COVID-19 infection. The dataset consisted of patient identifiers, demographic data, measured physiological parameters, other available clinical information and the outcome of the assessment (including whether the patient was conveyed to hospital).

Health and social care data relating to the population in England within the UK National Health Service (NHS) is managed by NHS Digital. We provided patient identifiers to NHS Digital to trace patients in our cohort and supply additional individual level demographic, co-morbidity and outcome data. NHS Digital identified records in their collections belonging to patients in our cohort, and provided data on patient demographics, limited COVID-related general practice (GP) records, emergency department attendances, hospital inpatient admissions, critical care periods, and death registrations from the UK Office of National Statistics.

YAS and NHS Digital removed records where patients indicated that they did not wish their data to be used for research purposes, via the NHS data opt-out service.^15^ The study team also excluded patients who had opted out of any part of the PRIEST study and those with inconsistent records (e.g. multiple deaths recorded or death before latest activity). Patient identifiers across all datasets were replaced with a consistent pseudo-identifier to enable the identification and linkage of records belonging to the same patient across all datasets but without revealing any patient’s identity.

### Inclusion Criteria

Our final cohort consisted of all adult (aged 16 years and over) patients at time of first (index) EMS attendance between 18^th^ March and 29^th^ June 2020, in which the attending ambulance staff recorded a clinical impression of suspected or confirmed COVID-19 infection, and who were successfully traced by NHS Digital.

### Outcome

The primary outcome was death, renal, respiratory, or cardiovascular organ support (identified from death registration and critical care data) at 30 days from index attendance.

The secondary outcome was death up to 30 days from index contact.

### Patient Characteristics

Physiological parameters were extracted from the first (primary) set of clinical observations recorded by the ambulance crew. Consistent with methods used to estimate the Charlson comorbidity index from the available routine data, comorbidities were included if recorded 12 months before the index EMS attendance.^16, 17^ In a similar way, only immunosuppressant drug prescriptions documented in GP records within 30 days before the index attendance, contributed to the immunosuppression co-morbidity variable. Pregnancy status was based on GP records recorded in the previous 9 months. Frailty in patients older than 65 years was derived from the latest recorded Clinical Frailty Scale (CFS) score (if recorded) in the electronic GP records prior to index attendance.^18^ Patients under the age of 65 years were not given a CFS score since it is not validated in this age group. However, if a CFS score was required to calculate a triage tool and the patient was under the age of 65, it was assumed to be 1. Performance status was estimated from the CFS.

### Analysis

We retrospectively applied the 5 triage tools to our cohort to assess their accuracy for the primary and secondary outcomes.^7, 10, 12-14^ Supplementary Material 1 provides details of scoring and handling missing data for the triage tools. For each tool we plotted the receiver operating characteristic (ROC) curve and calculated the area under the ROC curve (c-statistic) for discriminating between patients with and without adverse outcome. We calculated sensitivity, specificity, positive predictive value (PPV) and negative predictive value (NPV) at the following pre-specified decision making thresholds based on recommended or usual use: 0 vs 1+ CRB-65; 0–1 vs 2+ NEWS2; 0–2 vs 3+ PMEWS; 0–4 vs 5+ PRIEST; 0 vs 1 WHO score. The NEWS2 and PMEWS thresholds used are lower than previously proposed (0–3 vs 4+ NEWS and 0–3 vs 4+ PMEWS) for triaging patient acuity, and are based on the assessment of their performance in a UK ED population of patients with suspected COVID-19 infection, where higher thresholds gave sub-optimal sensitivity.^19^ The WHO algorithm and CRB-65 are positive if any criterion is positive. These tools were compared to the sensitivity, specificity, PPV and NPV of EMS clinicians’ decision to transfer patients to hospital. All analyses were based on assessment during the index EMS attendance and completed with SAS v9.4.

## Ethical Approval

The North West—Haydock Research Ethics Committee gave a favourable opinion on the PAINTED study on 25^th^ June 2012 (reference 12/NW/0303) and on the updated PRIEST study on 23rd March 2020, including the analysis presented here. The Confidentiality Advisory Group of the NHS Health Research Authority granted approval to collect data without patient consent in line with Section 251 of the National Health Service Act 2006. Access to data collected by NHS Digital was recommended for approval by its Independent Group Advising on the Release of Data (IGARD) on 11^th^ September 2021 having received additional recommendation for approval for access to GP records from the Profession Advisory Group (PAG) on 19^th^ August 2021.

## Patient Public Involvement

The Sheffield Emergency Care Forum (SECF) is a public representative group interested in emergency care research.^20^ Members of SECF advised on the development of the PRIEST study and two members joined the Study Steering Committee. Patients were not involved in the conduct of the study.

## Results

All totals presented from NHS Digital derived data sets (sex, number of current medications, comorbidities, clinical frailty scores and outcomes) are rounded to the nearest 5, with small numbers suppressed to comply with NHS Digital data disclosure guidance.

### Study population

Figure 1 and Table 1 summarise study cohort derivation and the characteristics of the 7,549 included individual adult patients. In total, 1,330 patients (17.6%, 95% CI:16.8% to 18.5%) experienced the primary outcome (death or organ support) and 1,065 (14.1%, 95% CI: 13.4% to 14.9%), the secondary outcome (death). Of the 7, 549 patients, the decision was made to transport 4,905 (65%) to hospital at index attendance. Of those, 1,120 (22.9%) experienced the primary adverse outcome. Of those not transported to hospital, 210 (7.9%) had an adverse outcome. Within the cohort, 3,925 patients (52%, 95% CI:50.9% to 53.1%) were admitted as inpatients and 2,785 (36.9%, 95% CI: 35.8% to 38%) had a diagnosis of COVID confirmed in hospital (since unrestricted community testing was not available until the 18/05/2020) within 30 days of index EMS attendance.

**Figure 1.**
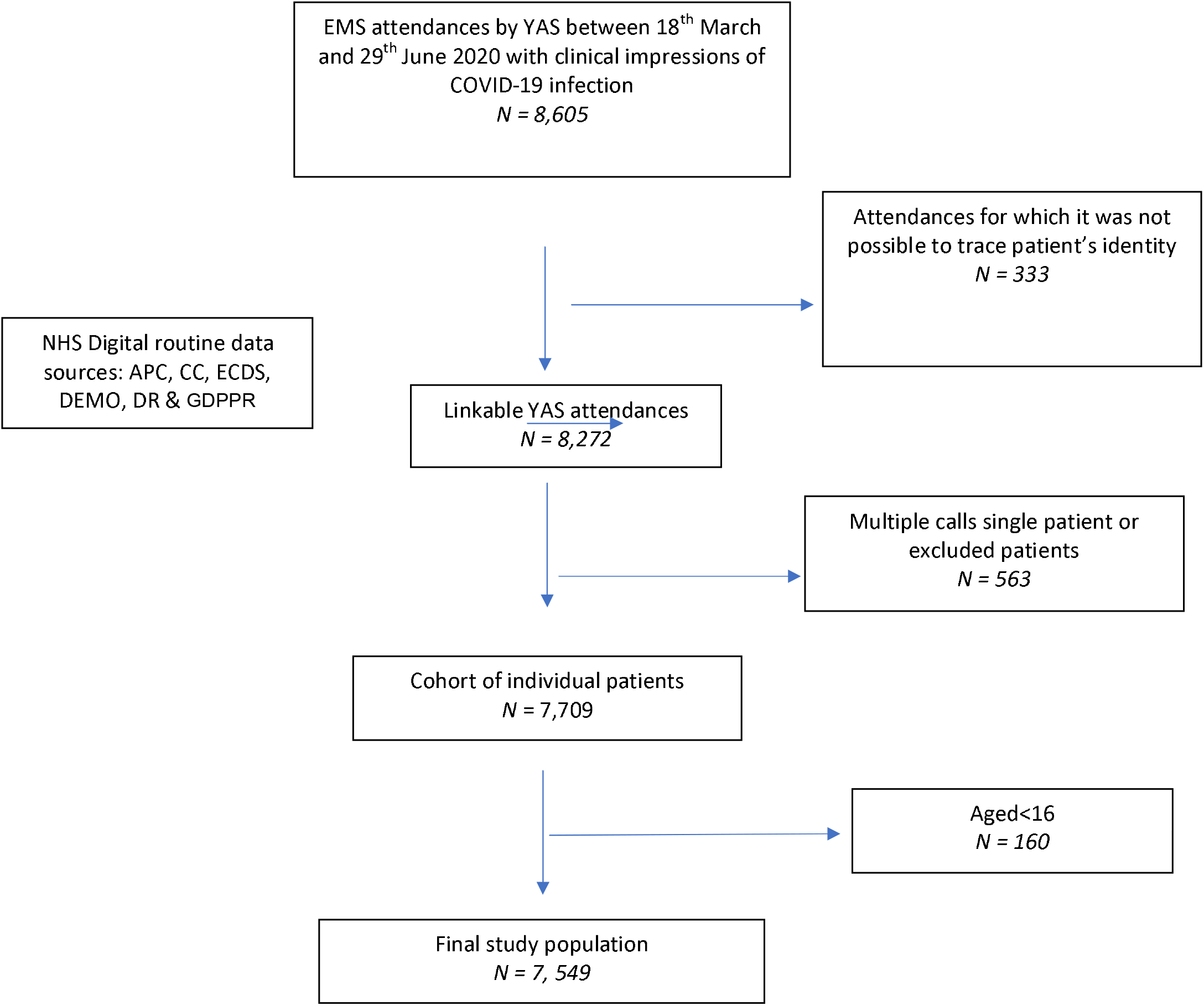
STROBE flow diagram of study population selection

**Table 1.**
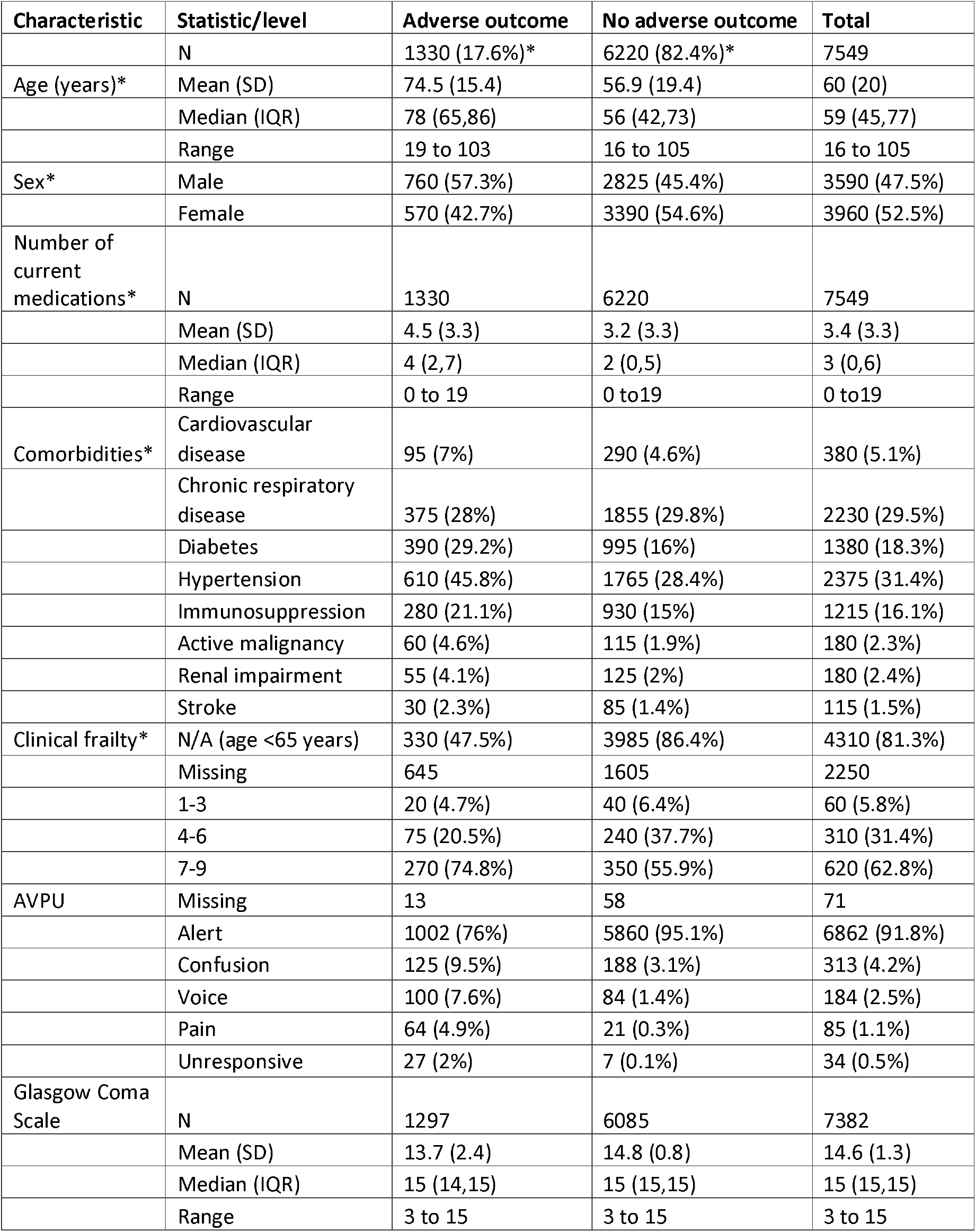
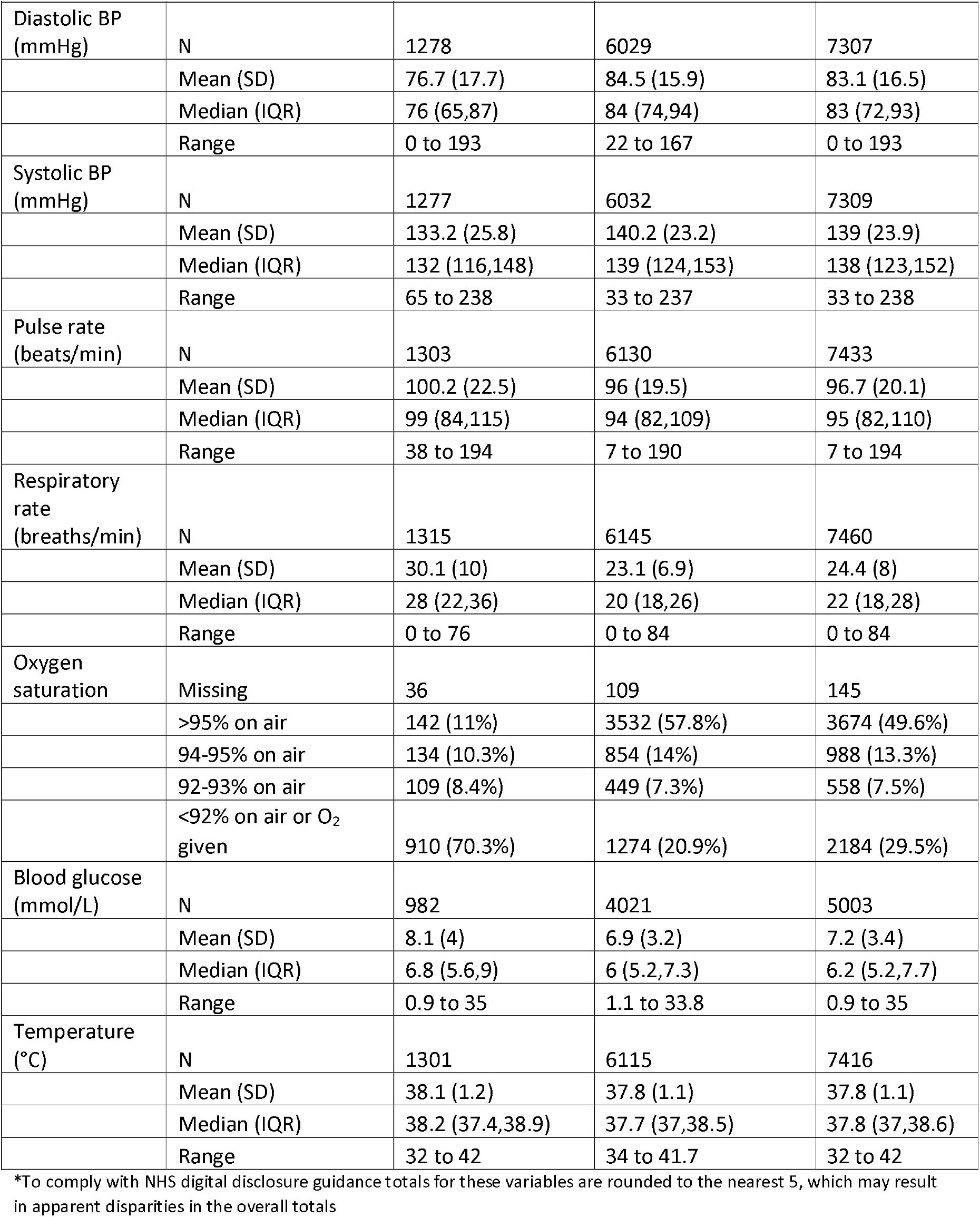
Patient characteristics by outcome

### Triage tool performance

Sensitivity, specificity, positive and negative predictive values for predicting the primary composite outcome using pre-defined score thresholds are provided in Table 2 and the secondary outcome of death in Table 3. Sensitivity and specificity statistics are provided for every score threshold in Supplementary Material 2. The ROC curves for these analyses are shown in Figures 2–3.

**Table 2.** Triage tool diagnostic accuracy statistics (95% CI) for predicting any adverse outcome

| Tool | N* | C-statistic | Threshold | Proportion with score | Sensitivity | Specificity | PPV | NPV |
| --- | --- | --- | --- | --- | --- | --- | --- | --- |
| CRB-65 | 7469 | 0.79<br>(0.78, 0.80) | >0 | 0.54 | 0.89<br>(0.88, 0.89) | 0.54<br>(0.53, 0.54) | 0.29 (0.29, 0.30) | 0.96 (0.95, 0.96) |
| NEWS2 | 7433 | 0.80<br>(0.78, 0.81) | >1 | 0.75 | 0.96<br>(0.96, 0.96) | 0.30<br>(0.29, 0.30) | 0.23 (0.22, 0.23) | 0.97 (0.97, 0.97) |
| PMEWS | 7460 | 0.81<br>(0.80, 0.83) | >2 | 0.72 | 0.98<br>(0.97, 0.98) | 0.34<br>(0.33, 0.34) | 0.24 (0.24, 0.24) | 0.99 (0.98, 0.99) |
| PRIEST | 7471 | 0.83<br>(0.82, 0.84) | >4 | 0.66 | 0.97<br>(0.97, 0.97) | 0.41<br>(0.40, 0.41) | 0.26 (0.25, 0.26) | 0.98 (0.98, 0.99) |
| WHO | 7471 | 0.64<br>(0.64, 0.65) | >0 | 0.74 | 0.98<br>(0.97, 0.98) | 0.31<br>(0.30, 0.31) | 0.23 (0.23, 0.24) | 0.98<br>(0.98, 0.99) |

\*Patients with 3 or more missing triage tool parameters were excluded from analysis when estimating performance

**Table 3.** Triage tool diagnostic accuracy statistics (95% CI) for predicting death within 30 days

| Tool | N* | C-statistic | Threshold | Proportion with score | Sensitivity | Specificity | PPV | NPV |
| --- | --- | --- | --- | --- | --- | --- | --- | --- |
| CRB-65 | 7469 | 0.84 (0.83, 0.85) | >0 | 0.54 | 0.95 (0.95, 0.96) | 0.53 (0.53, 0.54) | 0.25 (0.25, 0.26) | 0.99 (0.98, 0.99) |
| NEWS2 | 7433 | 0.78 (0.77, 0.80) | >1 | 0.75 | 0.95 (0.95, 0.96) | 0.28 (0.28, 0.29) | 0.18 (0.18, 0.18) | 0.97 (0.97, 0.98) |
| PMEWS | 7460 | 0.81 (0.80, 0.83) | >2 | 0.72 | 0.98 (0.98, 0.98) | 0.32 (0.32, 0.33) | 0.19 (0.19, 0.20) | 0.99 (0.99, 0.99) |
| PRIEST | 7471 | 0.85 (0.84, 0.86) | >4 | 0.66 | 0.98 (0.97, 0.98) | 0.39 (0.39, 0.40) | 0.21 (0.20, 0.21) | 0.99 (0.99, 0.99) |
| WHO | 7471 | 0.65 (0.64, 0.65) | >0 | 0.74 | 0.99 (0.99, 1.00) | 0.30 (0.30, 0.30) | 0.19 (0.19, 0.19) | 1.00 (1.00, 1.00) |

\*Patients with 3 or more missing triage tool parameters were excluded from analysis when estimating performance

**Figure 2.**
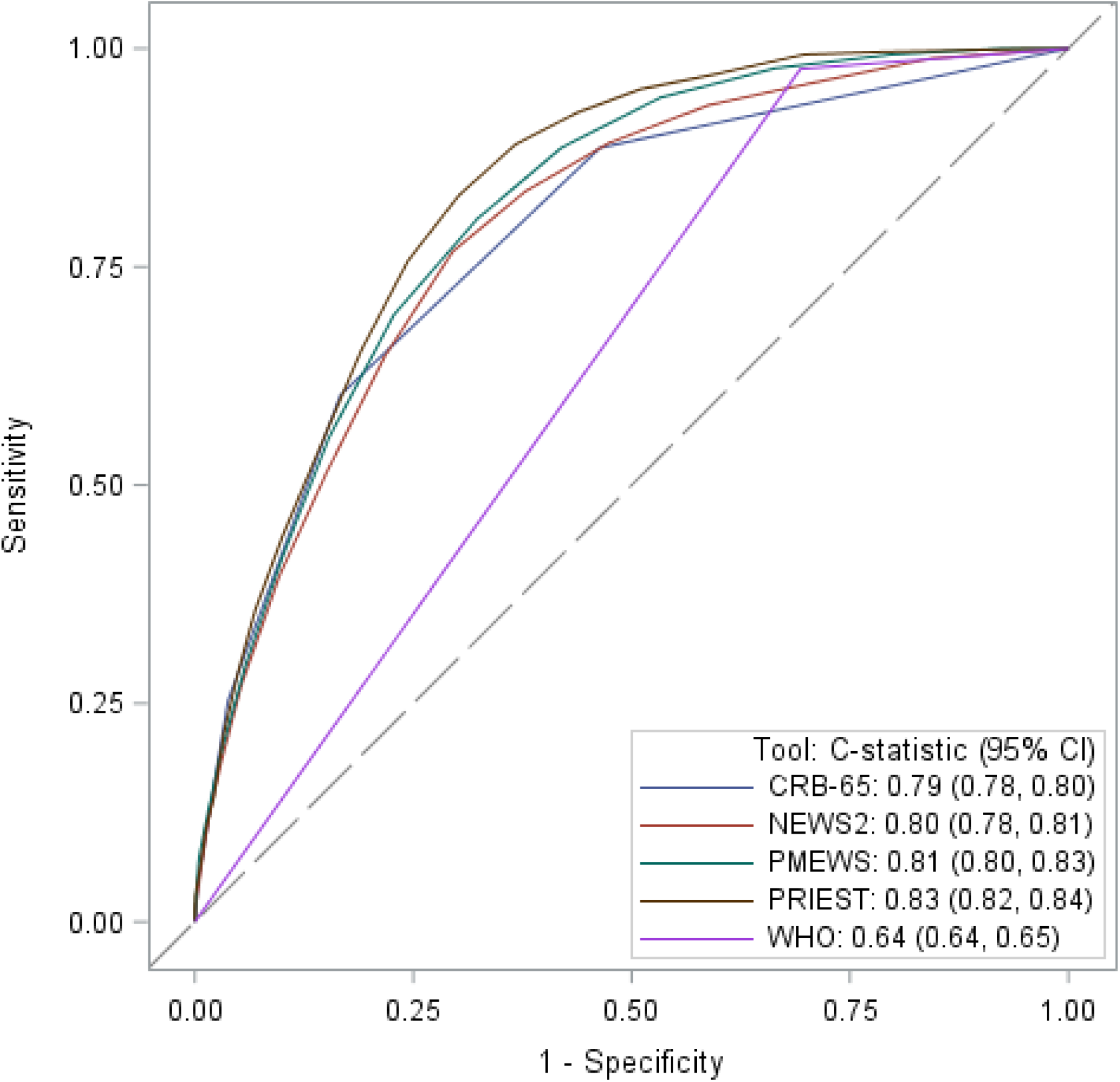
ROC curves showing triage tool performance for predicting any adverse outcome

**Figure 3.**
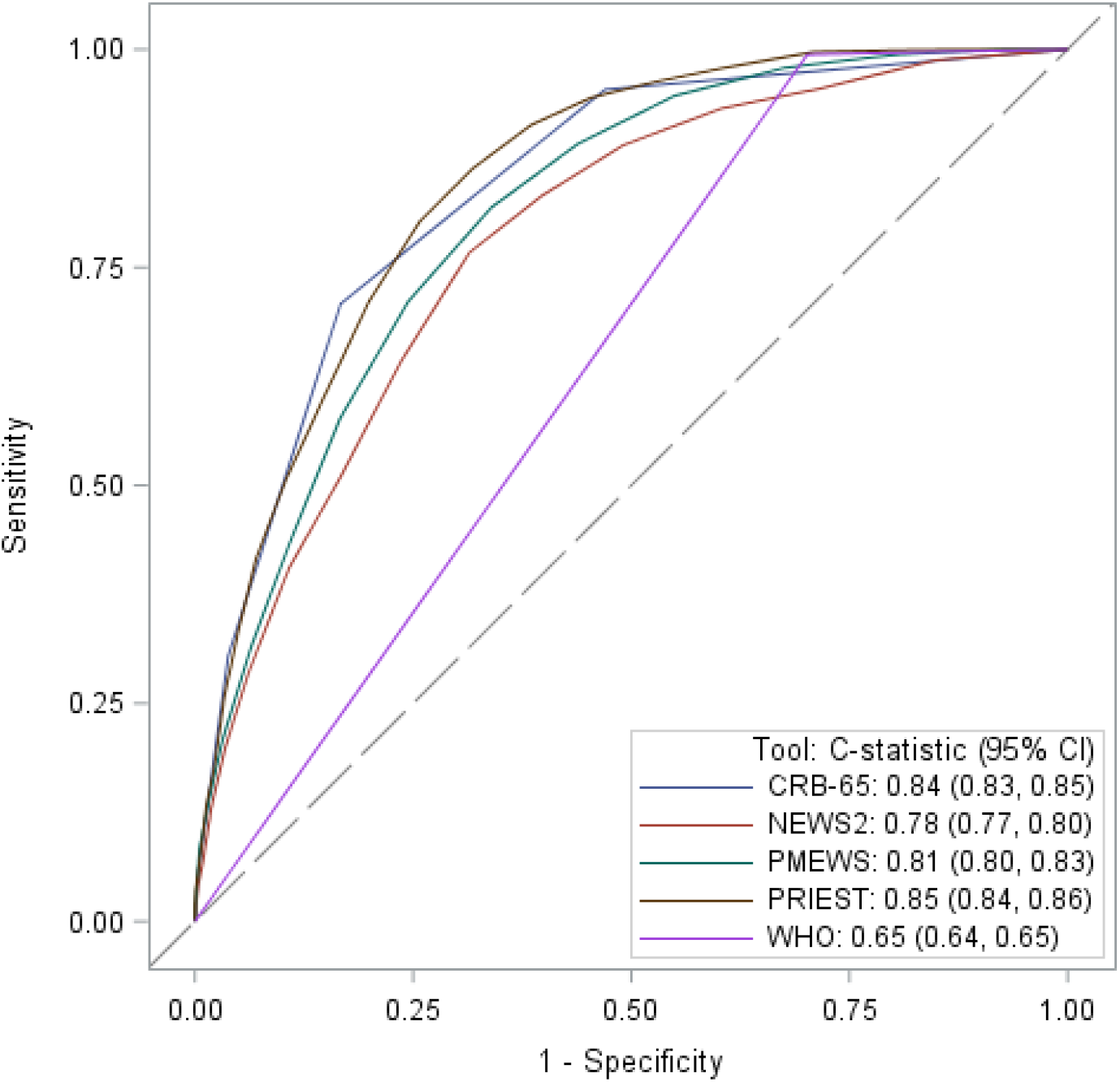
ROC curves showing triage tool performance for predicting death within 30 days

The PRIEST tool was robust to the removal of the performance status parameter; when doing so the C-statistic remained at 0.83 (95% CI 0.82 to 0.84). EMS decision to transfer patients to hospital had a sensitivity of 0.84 (95% CI 0.83 to 0.85) and specificity 0.39 (95% CI 0.39 to 0.40) for the primary outcome. The positive predictive value (PPV) was 0.23 (95% CI 0.22 to 0.23) and the negative predictive value (NPV) 0.92 (95% CI 0.92 to 0.92). Hypothetical use of any of the 5 triage tools would have achieved a higher sensitivity than the decision to transfer to hospital by the EMS crews within the cohort, but in the case of NEWS2, the WHO algorithm and PMEWS, this was at a cost of a lower specificity (Table 2). Of the tools assessed at the pre-determined thresholds, CRB-65 achieved the highest specificity but at the cost of sensitivity, and the PRIEST tool achieved a balance between both sensitivity, specificity and C-statistic of 0.83 (95% CI 0.82 to 0.84). The triage tools generally demonstrated better discrimination (except NEWS2) and a higher sensitivity for the secondary outcome, but a lower specificity (Table 3).

## Discussion

### Summary

The NEWS2, PMEWS, PRIEST tool and WHO algorithm identified patients at risk of adverse outcome with high sensitivity (>0.95) and specificity ranging between 0.3 (NEWS2) and 0.41 (PRIEST tool). They are therefore potentially suitable for use as triage tools to select patients for transfer to hospital. The high sensitivity of NEWS2 and PMEWS was achieved by using lower thresholds (NEWS2; 0–1 vs 2+ and PMEWS; 0–2 vs 3+) than previously recommended, based upon performance in an ED population of patients with suspected COVID-19 infection.^19^

At index attendance, 65% of patients were transported to hospital. Although a useful comparator for triage tool performance, the observed accuracy of EMS decision-making to transfer patients to hospital does not account for clinical best interest decisions not to covey patients to hospital who subsequently deteriorated, or patient wishes not to be conveyed. The sensitivity (0.84, 95% CI 0.83 to 0.85) and specificity (0.39, 95% CI 0.39 to 0.40) achieved by EMS decision making is nonetheless similar to that of tools used to triage undifferentiated patient acuity in the ED.^21^

To be clinically useful to EMS crews, the use of a triage tool would need to improve upon the existing sensitivity of clinical decision making, thereby reducing the risk of not transporting a patient to hospital who subsequently deteriorates, without leading to a disproportionately large increase in hospital conveyance. Use of any of the five triage tools at the pre-specified thresholds would potentially improve upon the sensitivity of existing EMS decision making. However, use of PMEWS, the WHO algorithm or NEWS2 would lead to up to a 10% increase in ED conveyances (Table 2). Use of both CRB65 and the PRIEST score would lead to improvements in sensitivity without sacrificing specificity. CRB65 achieved the highest specificity of any of the tools (0.54 95% CI: 0.53 to 0.54) and its use would reduce the number of patients conveyed to hospital by around 10%. However, patients not conveyed to hospital would have around a 4% risk of subsequently deteriorating. The PRIEST tool achieved a sensitivity of 0.97 (95% CI: 0.97 to 0.97) without increasing the number of patients transported to hospital. Using the PRIEST tool, patients who were not conveyed to hospital would have a 2% risk of subsequent deterioration (compared to an estimated 8% on EMS decision making in this cohort).

### Strengths and limitations

Previous evaluations of triage tool accuracy and prognostic COVID-19 prognostic research in the pre-hospital setting, are limited by only including patients who were subsequently admitted to hospital.^22-25^ This is the first evaluation to use a large cohort of patients identified from routinely collected EMS records and linked to nationally collected, patient-level, healthcare data to provide robust outcome data for all patients including those not conveyed to hospital. We had low rates of missing data in the variables used in the triage tools assessed (Table 1). We also assessed the performance of triage tools in a cohort of patients with suspected infection which, in the absence of accurate universally available rapid COVID-19 diagnostic tests, reflects the population which EMS staff must clinically triage. Most existing research either aimed to determine if patients with suspected infection have COVID-19, or to risk stratify patients with confirmed infection in a hospital setting.^26^

Our evaluation of triage tool accuracy is limited to a single ambulance service, albeit one covering a large population across the North of England, so the results may not be generalisable to other healthcare settings. Other ambulance services may serve populations with a different risk-profile, provide different types of EMS response or have different thresholds and guidelines regarding when to convey patients to hospital. The population used is likely to be have similar baseline characteristics to that used to derive and validate the PRIEST score in an ED population, as it was conducted at the same time at hospitals in the region (and elsewhere in the UK).^10^ A sensitivity analysis in which patients recruited to the ED-based PRIEST study were removed from analysis, did not affect estimates of triage tool performance. The PRIEST tool may perform less well if applied to a different, especially non-UK, health care setting.

We assumed that if co-morbidities were not recorded in routine data within the previous 12-months of the index event, they were not present. Our cohort is based on the clinical impression of likely COVID infection as determined by EMS crews. This is partly determined by prevalence of COVID-19 infection which varied during the study period however YAS guidance stated possible COVID-19 infection should be considered in all patients with shortness of breath, cough or fever and in patients with a history of close contact with someone with these symptoms.

### Implications

Clinical tools should be used in conjunction with clinical decision making when determining whether a patient needs to be conveyed to hospital by EMS crews. As previously highlighted, there may be good clinical reasons why patients who subsequently deteriorated were not conveyed to hospital in this cohort. Within this limitation, our study provides evidence that use of existing clinical triage tools may improve clinical decision making in a prehospital setting where the prevalence of serious adverse outcomes is similar to the ED.

In health care contexts where minimising risk of adverse outcomes in those not conveyed to hospital is the priority, use of PMEWS or the WHO criteria may be recommended, as they appear to optimise sensitivity. Use of the COVID-specific PRIEST tool would achieve almost the same gains in sensitivity (0.97 versus 0.98) without leading to a corresponding increase in patients being unnecessarily conveyed to hospital. The use of CRB65 would maximise specificity over gains in sensitivity, with a 4% risk of adverse outcomes in patients left to self-care in the community. This may be appropriate in resource constrained health care contexts and, as oxygen saturations do not form part of the assessment tool, it can be practically applied to a large range of health care settings.

Further research assessing triage tool performance alongside clinical judgement in the prehospital setting would be helpful to determine whether triage tools would improve accuracy of decisions to transfer patients to hospital in practice. Given the high prevalence of adverse outcomes in this cohort, the findings may not be applicable to other lower risk community settings (e.g. patients being assessed by general practitioners) and therefore similar research is needed for these populations.

## Conclusion

The NEWS2, PMEWS, PRIEST tool and WHO algorithm achieved high estimated sensitivities (>0.95) with respect to death or organ support, and specificities ranging between 0.3 (NEWS2) and 0.41 (PRIEST tool). EMS decision to transfer patients to hospital achieved a sensitivity of 0.84 (95% CI 0.83 to 0.85). Although, there may be good clinical reasons why patients who deteriorated were not conveyed to hospital, use of these triage tools would potentially improve EMS triage of patients with suspected COVID-19 infection. Use of the PRIEST tool would lead to significant gains in sensitivity without increasing the number of patients conveyed to hospital.

## Data Availability

The data used for this study are subject to data sharing agreements with NHS Digital and Yorkshire
Ambulance Service which prohibits further sharing of individual level data. The data sets used are
obtainable from these organisations subject to necessary authorisations and approvals.

