## Supplementary Material for "Prognostic accuracy of triage tools for adults with suspected COVID-19 in a pre-hospital setting: an observational cohort study"

**Supplementary Material 1: Triage tool scoring details**

**CRB-65:**

The CRB-65 score uses four parameters, each scoring 1 point when positive and zero if negative, to give a total score between zero and five.

Four parameters:

1. Confusion: GCS-V is less than 4 or GCS total is less than 15 or AVPU is recorded as V, P or U
2. Respiratory: rate of 30 breaths per minute or more
3. Blood pressure: diastolic BP is 60mmHg or less or systolic BP is 90 mmHg or less
4. Age: 65 years or more

Missing data: The rules above effectively classify missing data as normal. The CRB-65 score is recommended for community settings where access to blood testing is more limited. It is a 4-point scale that does not include urea with the threshold as <2 for low risk, 2+ for high risk. If a patient has fewer than three of the five parameters complete, the score was not be calculated.

**PMEWS:**

PMEWS uses six physiological parameters and patient parameters to calculate a score from zero to 19. The score is calculated by taking the score in the table below dependent on each of the six physiological parameters then adding points for two patient parameters after if they are positive.

Physiological:

| **Score** | **3** | **2** | **1** | **0** | **1** | **2** | **3** |
| --- | --- | --- | --- | --- | --- | --- | --- |
| **Respiratory Rate** | ≤8 |  |  | 9-18 | 19-25 | 26-29 | ≥30 |
| **SaO₂** | <89 | 90-93 | 94-96 | >96 |  |  |  |
| **Pulse Rate** | ≤40 | 41-50 |  | 51-100 | 101-110 | 111-129 | ≥130 |
| **Systolic BP** | ≤70 | 71-90 | 90-100 | >100 |  |  |  |
| **Temperature** |  | ≤35.0 | 35.1-36.0 | 36.1-37.9 | 38-38.9 | ≥39 |  |
| **Neuro** |  |  |  | Alert | Confused Agitated* | Voice | Pain Uncon |

* confused/agitated will be defined based on GCS-V<4 or GCS total<15

Patient:

1. Add 1 point if age>65
2. Add 1 point if either:
   a. Patient lives alone / no fixed abode or
   b. has a co-morbidity (respiratory, cardiac, renal, immunosuppressed, diabetes)
   c. performance status is more than two suggesting limited activity can self-care, limited activity limited self-care, or bed/chair bound no self-care.

Missing data: If data is missing one or two variables then the normal score (zero) was assumed. If more than three variables are missing the patient was excluded. These rules effectively classify missing data as normal. If AVPU was missing and GCS was recorded, impute the following AVPU scores using GCS: 0 if GCS=15, 1 if GCS=12-14; 2 if GCS=9-11 and 3 if GCS<9.

**NEWS2:**

The NEWS2 has seven parameters which are scores from zero to three providing an overall score between zero and 20. The scores for each parameter can be found in the table below.

| **Score** | **3** | **2** | **1** | **0** | **1** | **2** | **3** |
| --- | --- | --- | --- | --- | --- | --- | --- |
| **Respiratory Rate** | ≤8 |  | 9-11 | 12-20 |  | 21-24 | ≥25 |
| **SaO₂** | ≤91 | 92-93 | 94-95 | ≥96 |  |  |  |
| **Pulse Rate** | ≤40 |  | 41-50 | 51-90 | 91-110 | 111-130 | ≥131 |
| **Systolic BP** | ≤90 | 91-100 | 101-110 | 111-219 |  |  | ≥220 |
| **Temperature** | ≤35.0 |  | 35.1-36.0 | 36.1-38.0 | 38.1-39.0 | ≥39.1 |  |
| **Neuro** |  |  |  | Alert |  |  | Confusion, Voice, Pain, Unresponsive |
| **Air or Oxygen** |  | Oxygen (based on FiO_2_>21%, or FiO_2_>0 L/min) |  | Air |  |  |  |

Missing data: Any missing data will be imputed with the value zero, therefore classifying missing as normal. The score was calculated if fewer than three of the parameters were available.

**WHO decision making algorithm for hospitalisation with pneumonia:**

The WHO decision making algorithm for hospitalisation with pneumonia suggests an adult patient is admitted (score 1) if any of the following are present:

- respiratory rate >30/minute,
- oxygen saturation <90%,
- respiratory distress (not included in this evaluation),
- age >60,
- any of the following comorbidities; hypertension, diabetes, cardiovascular disease, chronic respiratory disease, renal impairment or immunosuppression

As a subjective clinical assessment of respiratory distress is not routinely consistently recorded in our data this was not be included. Any missing data was assumed as normal. A score was not calculated if fewer than three of the above were complete.

**The PRIEST clinical severity Score:**

The core PRIEST clinical severity score consists of the seven parameters of NEWS2 and age, sex and performance status. We will assessed the full score and a version which omits performances status as this may not be reliably available from GP records. Scores for each parameter are as follows.

| **Variable** | **Range** | **Score** |
| --- | --- | --- |
| Respiratory rate (per minute) | 12-20 | 0 |
|  | 9-11 | 1 |
|  | 21-24 | 2 |
|  | <9 or >24 | 3 |
| Oxygen saturation (%) | >95 | 0 |
|  | 94-95 | 1 |
|  | 92-93 | 2 |
|  | <92 | 3 |
| Heart rate (per minute) | 51-90 | 0 |
|  | 41-50 or 91-110 | 1 |
|  | 111-130 | 2 |
|  | <41 or >130 | 3 |
| Systolic BP (mmHg) | 111-219 | 0 |
|  | 101-110 | 1 |
|  | 91-100 | 2 |
|  | <91 or >219 | 3 |
| Temperature (C) | 36.1-38.0 | 0 |
|  | 35.1-36.0 or 38.1-39.0 | 1 |
|  | >39.0 | 2 |
|  | <35.1 | 3 |
| Alertness | Alert | 0 |
|  | Confused or not alert | 3 |
| Inspired oxygen | Air | 0 |
|  | Supplemental oxygen | 2 |
| Sex | Female | 0 |
|  | Male | 1 |
| Age (years) | 16-49 | 0 |
|  | 50-65 | 2 |
|  | 66-80 | 3 |
|  | >80 | 4 |
| Performance status | Unrestricted normal activity | 0 |
|  | Limited strenuous activity, can do light activity | 1 |
|  | Limited activity, can self-care | 2 |
|  | Limited self-care | 3 |
|  | Bed/chair bound, no self-care | 4 |

Missing data in NEWS2 parameters was handled in the same way as for NEWS2. Performance status was be assumed normal if a clinical frailty scale has not been completed in linked GP records

**Supplementary Material 2: Performance of triage tools across the whole range of available scores**

|  |  | **Primary: any adverse outcome** | | **Secondary: death** | |
| --- | --- | --- | --- | --- | --- |
| **Tool** | **Threshold** | **Sensitivity** | **Specificity** | **Sensitivity** | **Specificity** |
| CRB-65 | >0 | 0.89 (0.88,0.89) | 0.54 (0.53,0.54) | 0.95 (0.95,0.96) | 0.53 (0.53,0.54) |
|  | >1 | 0.6 (0.59,0.61) | 0.83 (0.83,0.84) | 0.71 (0.7,0.72) | 0.83 (0.83,0.84) |
|  | >2 | 0.25 (0.24,0.26) | 0.96 (0.96,0.96) | 0.3 (0.29,0.32) | 0.96 (0.96,0.96) |
|  | >3 | 0.05 (0.04,0.05) | 1 (1,1) | 0.06 (0.05,0.06) | 1 (1,1) |
| NEWS2 | >0 | 0.99 (0.99,0.99) | 0.16 (0.16,0.16) | 0.99 (0.98,0.99) | 0.15 (0.15,0.16) |
|  | >1 | 0.96 (0.96,0.96) | 0.3 (0.29,0.3) | 0.95 (0.95,0.96) | 0.28 (0.28,0.29) |
|  | >2 | 0.93 (0.93,0.94) | 0.41 (0.41,0.42) | 0.93 (0.93,0.94) | 0.4 (0.39,0.4) |
|  | >3 | 0.89 (0.88,0.9) | 0.53 (0.52,0.53) | 0.89 (0.88,0.9) | 0.51 (0.5,0.51) |
|  | >4 | 0.84 (0.83,0.84) | 0.62 (0.62,0.63) | 0.83 (0.82,0.84) | 0.6 (0.6,0.61) |
|  | >5 | 0.77 (0.76,0.77) | 0.7 (0.7,0.71) | 0.77 (0.76,0.78) | 0.69 (0.68,0.69) |
|  | >6 | 0.65 (0.64,0.66) | 0.78 (0.78,0.79) | 0.64 (0.63,0.65) | 0.76 (0.76,0.77) |
|  | >7 | 0.51 (0.5,0.52) | 0.85 (0.85,0.85) | 0.51 (0.5,0.52) | 0.83 (0.83,0.84) |
|  | >8 | 0.39 (0.38,0.4) | 0.9 (0.9,0.91) | 0.4 (0.39,0.41) | 0.89 (0.89,0.9) |
|  | >9 | 0.27 (0.26,0.28) | 0.95 (0.94,0.95) | 0.28 (0.27,0.29) | 0.94 (0.94,0.94) |
|  | >10 | 0.18 (0.18,0.19) | 0.97 (0.97,0.97) | 0.2 (0.19,0.21) | 0.97 (0.96,0.97) |
|  | >11 | 0.12 (0.11,0.13) | 0.98 (0.98,0.98) | 0.13 (0.12,0.14) | 0.98 (0.98,0.98) |
|  | >12 | 0.06 (0.06,0.07) | 0.99 (0.99,0.99) | 0.07 (0.06,0.07) | 0.99 (0.99,0.99) |
|  | >13 | 0.04 (0.03,0.04) | 1 (1,1) | 0.04 (0.04,0.05) | 1 (1,1) |
|  | >14 | 0.02 (0.01,0.02) | 1 (1,1) | 0.02 (0.01,0.02) | 1 (1,1) |
|  | >15 | 0.01 (0.01,0.01) | 1 (1,1) | 0.01 (0.01,0.01) | 1 (1,1) |
|  | >16 | 0 (0,0) | 1 (1,1) | 0 (0,0.01) | 1 (1,1) |
|  | >17 | 0 (0,0) | 1 (1,1) | 0 (0,0) | 1 (1,1) |
| PMEWS | >0 | 1 (1,1) | 0.08 (0.07,0.08) | 1 (1,1) | 0.07 (0.07,0.08) |
|  | >1 | 0.99 (0.99,0.99) | 0.2 (0.2,0.21) | 0.99 (0.99,1) | 0.2 (0.19,0.2) |
|  | >2 | 0.98 (0.97,0.98) | 0.34 (0.33,0.34) | 0.98 (0.98,0.98) | 0.33 (0.32,0.33) |
|  | >3 | 0.94 (0.94,0.95) | 0.47 (0.46,0.47) | 0.95 (0.94,0.95) | 0.45 (0.45,0.46) |
|  | >4 | 0.89 (0.88,0.89) | 0.58 (0.58,0.59) | 0.89 (0.88,0.9) | 0.56 (0.56,0.57) |
|  | >5 | 0.8 (0.8,0.81) | 0.68 (0.67,0.68) | 0.82 (0.81,0.83) | 0.66 (0.66,0.66) |
|  | >6 | 0.69 (0.69,0.7) | 0.77 (0.77,0.78) | 0.71 (0.7,0.72) | 0.76 (0.75,0.76) |
|  | >7 | 0.55 (0.54,0.56) | 0.85 (0.84,0.85) | 0.57 (0.56,0.59) | 0.83 (0.83,0.84) |
|  | >8 | 0.41 (0.4,0.42) | 0.9 (0.9,0.9) | 0.43 (0.42,0.44) | 0.89 (0.89,0.89) |
|  | >9 | 0.29 (0.29,0.3) | 0.94 (0.94,0.94) | 0.31 (0.3,0.32) | 0.94 (0.93,0.94) |
|  | >10 | 0.19 (0.18,0.19) | 0.97 (0.97,0.97) | 0.21 (0.2,0.22) | 0.97 (0.97,0.97) |
|  | >11 | 0.11 (0.11,0.12) | 0.99 (0.99,0.99) | 0.13 (0.12,0.14) | 0.99 (0.99,0.99) |
|  | >12 | 0.07 (0.07,0.08) | 1 (1,1) | 0.08 (0.08,0.09) | 1 (0.99,1) |
|  | >13 | 0.04 (0.03,0.04) | 1 (1,1) | 0.04 (0.04,0.05) | 1 (1,1) |
|  | >14 | 0.01 (0.01,0.02) | 1 (1,1) | 0.02 (0.01,0.02) | 1 (1,1) |
|  | >15 | 0.01 (0.01,0.01) | 1 (1,1) | 0.01 (0.01,0.01) | 1 (1,1) |
|  | >16 | 0 (0,0) | 1 (1,1) | 0 (0,0) | 1 (1,1) |
| PRIEST | >0 | 1 (1,1) | 0.05 (0.05,0.05) | 1 (1,1) | 0.05 (0.05,0.05) |
|  | >1 | 1 (1,1) | 0.12 (0.12,0.13) | 1 (1,1) | 0.12 (0.12,0.12) |
|  | >2 | 1 (0.99,1) | 0.21 (0.21,0.22) | 1 (1,1) | 0.21 (0.2,0.21) |
|  | >3 | 0.99 (0.99,0.99) | 0.31 (0.3,0.31) | 0.99 (0.99,1) | 0.29 (0.29,0.3) |
|  | >4 | 0.97 (0.97,0.97) | 0.41 (0.4,0.41) | 0.98 (0.97,0.98) | 0.39 (0.39,0.4) |
|  | >5 | 0.95 (0.95,0.95) | 0.49 (0.49,0.5) | 0.96 (0.95,0.96) | 0.48 (0.47,0.48) |
|  | >6 | 0.92 (0.92,0.93) | 0.57 (0.56,0.57) | 0.94 (0.93,0.95) | 0.55 (0.55,0.55) |
|  | >7 | 0.89 (0.88,0.89) | 0.64 (0.63,0.64) | 0.91 (0.9,0.92) | 0.62 (0.62,0.62) |
|  | >8 | 0.83 (0.82,0.83) | 0.7 (0.7,0.71) | 0.86 (0.85,0.87) | 0.69 (0.68,0.69) |
|  | >9 | 0.75 (0.74,0.76) | 0.76 (0.76,0.76) | 0.8 (0.79,0.81) | 0.75 (0.74,0.75) |
|  | >10 | 0.65 (0.64,0.66) | 0.81 (0.81,0.82) | 0.7 (0.69,0.71) | 0.8 (0.8,0.81) |
|  | >11 | 0.53 (0.52,0.54) | 0.86 (0.86,0.87) | 0.59 (0.58,0.6) | 0.86 (0.85,0.86) |
|  | >12 | 0.44 (0.43,0.44) | 0.9 (0.9,0.9) | 0.5 (0.49,0.51) | 0.9 (0.9,0.9) |
|  | >13 | 0.35 (0.34,0.36) | 0.93 (0.93,0.94) | 0.41 (0.4,0.42) | 0.93 (0.93,0.93) |
|  | >14 | 0.28 (0.27,0.28) | 0.95 (0.95,0.96) | 0.33 (0.32,0.34) | 0.95 (0.95,0.95) |
|  | >15 | 0.21 (0.2,0.22) | 0.97 (0.97,0.97) | 0.25 (0.24,0.26) | 0.97 (0.97,0.97) |
|  | >16 | 0.14 (0.14,0.15) | 0.98 (0.98,0.98) | 0.17 (0.17,0.18) | 0.98 (0.98,0.98) |
|  | >17 | 0.11 (0.1,0.11) | 0.99 (0.99,0.99) | 0.13 (0.12,0.14) | 0.99 (0.99,0.99) |
|  | >18 | 0.07 (0.06,0.07) | 0.99 (0.99,0.99) | 0.08 (0.08,0.09) | 0.99 (0.99,0.99) |
|  | >19 | 0.04 (0.04,0.05) | 1 (1,1) | 0.05 (0.05,0.06) | 1 (1,1) |
|  | >20 | 0.03 (0.02,0.03) | 1 (1,1) | 0.03 (0.03,0.04) | 1 (1,1) |
|  | >21 | 0.02 (0.01,0.02) | 1 (1,1) | 0.02 (0.02,0.02) | 1 (1,1) |
|  | >22 | 0.01 (0.01,0.01) | 1 (1,1) | 0.01 (0.01,0.01) | 1 (1,1) |
|  | >23 | 0 (0,0.01) | 1 (1,1) | 0 (0,0.01) | 1 (1,1) |
|  | >24 | 0 (0,0) | 1 (1,1) | 0 (0,0) | 1 (1,1) |
|  | >25 | 0 (0,0) | 1 (1,1) | 0 (0,0) | 1 (1,1) |
| WHO | >0 | 0.98 (0.97,0.98) | 0.31 (0.3,0.31) | 0.99 (0.99,1) | 0.3 (0.3,0.3) |
